# Association of Shared Care Networks with Heart Failure Excessive Hospital Readmissions

**DOI:** 10.1101/2021.04.07.21255061

**Authors:** Diego Pinheiro, Ryan Hartman, Jing Mai, Erick Romero, Saad Soroya, Carmelo Bastos-Filho, Ricardo Lima, Michael Gibson, Imo Ebong, Julie T. Bidwell, Miriam Nuño, Martin Cadeiras

## Abstract

**Objectives:** This study aimed to evaluate the impact of shared care networks on heart failure readmission rates.

**Background:** Higher-than-expected heart failure (HF) readmissions affect half of US hospitals every year. The Hospital Reduction Readmission Program (HRRP) has reduced risk-adjusted readmissions, but it has also produced unintended consequences. Shared care models have been advocated for HF care, but the association of shared care networks with HF readmissions has never been investigated.

**Methods:** We curated publicly available data on hospital discharges and HF excessive readmission ratios (ERRs) from hospitals in California between 2012 and 2017. Shared Care Areas (SCAs) were delineated as data-driven units of care coordination emerging from discharge networks. The localization index (LI), the proportion of patients who reside in the same SCA in which they are admitted, were calculated by year. Generalized estimating equations (GEE) were used to evaluate the association between the LI and the ERR of hospitals controlling for race/ethnicity and socioeconomics factors.

**Results:** A total of 300 hospitals in California in a 6-yr period were included. The HF excessive readmission ratio (ERR) was negatively associated with the localization index (beta: -0.0474; 95% CI: -0.082 to -0.013). The percentage of Black residents within the SCAs was the only statistically significant covariate (beta: 0.4128; 95% CI: 0.302 to 0.524).

**Conclusions:** Higher-than-expected HF readmissions were associated with shared care networks. Control mechanisms such as the HRRP may need to characterize and reward shared care to guide hospitals towards a more organized HF care system.

## INTRODUCTION

Higher-than-expected heart failure (HF) readmission impacts approximately half of US hospitals every year, and almost every hospital has experienced it at least once in the period between 2012 and 2017 (Table 1). By 2030, HF is projected to affect at least 8 million people in the US, with an incidence of 21 per 1000 people over 65 years of age, and estimated cost of $69.8 billion (2). The number of HF patients receiving HF care and requiring advanced HF therapies such as left ventricular assisted devices (LVAD) will grow exponentially (3). Addressing higher-than-expected HF readmissions for HR patients as demand increases with the aging population require improved care coordination mechanisms that promote a more organized HF care system (4).

**Table 1.** Descriptive Statistics of Excessive Readmission Ratio (ERR) and Percentage of Hospitals Penalized in the US and California.

|  |  |  |  |  |  |  |
| --- | --- | --- | --- | --- | --- | --- |
|  | 2012 | 2013 | 2014 | 2015 | 2016 | 2017 |
| United States |  |  |  |  |  |  |
|  |  |  |  |  | 2,82 | 2,79 |
| Hospitals, N | 2,864 | 2,860 | 2,825 | 2,820 | 7 | 3 |
|  |  |  |  |  | 49.4 | 48.9 |
| Hospitals Penalized, % | 49.76 | 48.95 | 49.17 | 49.22 | 5 | 4 |
| Excessive Readmission Ratio |  |  |  |  | 1.00 | 1.00 |
| (ERR) | 1.0013 | 1.0012 | 1.0010 | 1.0012 | 18 | 16 |
|  |  |  |  |  | 0.07 | 0.07 |
| ERR, std | 0.0844 | 0.0809 | 0.0803 | 0.0774 | 76 | 53 |
| California |  |  |  |  |  |  |
| Hospitals, N | 233 | 233 | 233 | 233 | 237 | 237 |
|  |  |  |  |  | 51.9 | 56.1 |
| Hospitals Penalized, % | 49.36 | 48.50 | 56.22 | 55.79 | 0 | 2 |
| Excessive Readmission Ratio |  |  |  |  | 1.00 | 1.00 |
| (ERR) | 0.9914 | 0.9963 | 1.0034 | 1.0057 | 49 | 87 |
|  |  |  |  |  | 0.07 | 0.07 |
| ERR, std | 0.0761 | 0.0778 | 0.0760 | 0.0731 | 20 | 03 |

HF is managed through a complex system that serves both affluent and vulnerable patient populations and encompasses nonlinear interactions among primary care, general cardiology, specialized HF clinics, and tertiary and quaternary centers. The implementation of any control mechanism can produce unintended consequences if the complexity of the HF care system is not taken into consideration (5)(6). Systemwide control programs such as the Hospital Reduction Readmission Program (HRRP) (7) may be a first step towards organizing the HF care system. Nonetheless, they will continue to create unintended consequences and penalize hospitals for factors beyond their control (8) unless these programs specifically foster care coordination mechanisms capable of promoting organization for HF care’s complex system.

Shared care integrates primary, secondary, and tertiary levels of care (9) and has been advocated as a necessary model to promote a more organized HF care system (10) such as the spoke-hub-and-node model (11). Shared care has been studied among chronic diseases (12), including HF (13), but only recently it has been advocated by international working groups as a way to organize HF care (10), particularly among advanced HF patients (11) such as patients with left ventricular assist device (LVAD) support (14). Shared Care Areas (SCAs) are data-driven units of care coordination captured from large-scale data on hospital discharges to patient residencies and SCAs may explain variation in medical adherence to HF guideline-directed medical therapy (15). The localization index of a SCA is the proportion of patients who reside in the same SCA they are admitted, and is a measure of local care coordination commonly used to evaluate SCAs (16). This study aimed to evaluate the longitudinal association between higher-than-expected HF readmissions and the localization index of SCAs both unadjusted and adjusted for racial/ethnic and socioeconomic factors.

## METHODS

### Study Population and Design

This retrospective longitudinal design included all hospitals from California that were eligible in the Hospital Readmissions Reduction Program (HRRP) from 2012 to 2017 (7) for which discharge data were also made publicly available from the Office of Statewide Health Planning and Development (OSHPD) (17). Hospitals with less than 2 repeated measures of higher-than-expected HF readmission in the HRRP or without discharge data in the OSHPD were excluded. Between 233 and 237 hospitals were included depending on the year. Ethical approval was unnecessary because all data is already made publicly available from both HRRP and OSHPD. All of the code, curated data, built networks, and data analysis resulting from this work are available on the Open Science Framework (OSF) repository of this work^1^.

### Data Sources

This study used Excessive Readmission Ratio (ERR) and Patient Origin/Market Share data made publicly available from the Hospital Reduction Readmission Program (HRRP) (7) and the Office of Statewide Health Planning and Development (OSHPD) (17), respectively. The ERR is a risk-standardized 30-day readmission ratio that adjusts for a set of patient-specific covariates such as congestive HF, renal failure, and chronic obstructive pulmonary disease (18). The ERR data of each year in the period from 2012 to 2017 (i.e., Fiscal Year [FY] 2014 and 2019) was separately downloaded from HRRP and compiled into a single file. Similarly, the Patient Origin/Market Share data are aggregated numbers of emergency department (ED) discharges among ZIP Codes of hospitals and patient residencies. ZIP Codes were converted to the respective ZIP Code Tabulation Area (ZCTAs) (19).

### Uncovering Shared Care Areas and Localization Index from Hospital-Patient Discharge Data

Six yearly hospital-patient discharge networks were built from OSHPD hospital-patient ED discharges between 2012 to 2017. In a hospital-patient discharge network (16), a node is the ZCTA of a hospital or patient residency, and the link between two nodes (i.e., ZCTAs) is the total number of ED discharges. For each yearly hospital-patient discharge network, shared care areas (SCAs) were delineated using community detection algorithms. Each delineated SCA consists of a set of ZCTAs in which hospitals are embedded. A set of four diverse community detection algorithms were considered to decrease both variability and bias (20). The algorithms were Louvain (21) with resolution equals to 1, Stochastic Block Model (22)(23) with degree corrected, Infomap (24) with two levels, and Speaker-Listener Label Propagation (25) with post-processing threshold equals to 0.5. The localization index (LI) represents the proportion of patient discharges from hospitals within the same SCA of which these patients live (26) (27). A higher localization index represents a homogenous SCA with localized care coordination (i.e., patients tend to receive care where they live).

### Statistical Analysis

The ERR hospitals and the localization index of SCAs were integrated at each year by linking the ZCTAs of hospitals and SCAs (Supplemental Table 1 and Supplemental Figure 1). A longitudinal regression was specified in which the dependent variable ERR of a hospital at time *t* as a function of the localization index of its SCA at time *t*. We used a generalized estimating equation (GEE) with an exchangeable correlation structure to account for multiple observations of ERR from the same hospital across years and SCAs (28). The estimated regression coefficients (beta) were used to measure unadjusted associations between the dependent and independent variables and adjusted associations after controlling for racial-ethnic and socioeconomics confounders associated with HF readmission at the regional level (29). The GEE was estimated using the Statsmodels python package (30). Also, hospitals were stratified based on quartiles of localization index and all covariates that were found statistically significant, and median values of ERRs and percentage of hospitals penalized were calculated for each quartile (Q1, Q2, Q3, Q4). We estimated 95% confidence intervals using 10,000 bootstrap samples with replacement from each quartile-the estimation of confidence intervals of medians using the Bootstrapped python package (31).

### Predicting Higher-Than-Expected Heart Failure Readmissions for Changes in Localization Index

The estimated GEE model was used to predict HF’s excessive readmission ratios (ERR) assuming a range of changes in the localization index in SCAs with distinct percentages of Black residents, the only statistically significant covariate. The differences in localization index between subsequent years were calculated for all hospitals. The 25^th^, 50^th^, and 75^th^ percentiles were separately calculated for both positive (+q1, +q2, and +q3) and negative (−q1, -q2, -q3) differences. The SCAs were stratified by quartiles of Black residents (Q1, Q2, Q3, and Q4). The ERR was predicted using the GEE model after each positive and negative percentile difference in localization index was applied to the stratified SCA data.

## RESULTS

### Descriptive Statistics of Heart Failure Hospital Readmissions in the United States and California

The ERR is calculated every year by the HRRP for the approximately 2.7 to 2.9 thousand hospitals in the US, from which 233 to 237 hospitals are from California (Table 1). Overall, approximately half of US hospitals are penalized, and this percentage has not changed during the study period between 2012 to 2017. The ERR (and the percentage of hospitals penalized) of US hospitals have remained approximately constant during the study period, from 1.0013 (49.76%) in 2012 to 1.0016 (48.94%) in 2017. The ERR (and the percentage of hospitals penalized) of hospitals in California increased from 0.9914 (49.36%) to 1.0087 (56.12%). In 2017, the percentage of hospitals penalized in California 56.12% (95% CI, 49.75% to 62.29%) is slightly higher than that among all hospitals in the US 48.91% (95% CI, 47.06% to 50.76%). Although not statistically significant, the ERR standard deviation appears to be decreasing over the years.

### Association of Excessive Readmission Ratio and Localization Index

The results of the regression analysis (Figure 1 and Table 2) indicate that the ERR of hospitals was negatively associated with the localization index of their SCAs (e.g. ERRs were lower when hospitals were located in SCAs where more patients received care close to where they resided) according to both unadjusted (beta = -0.0717; *p*<.001) and adjusted (beta = -0.0495; *p*= p=0.0495) coefficients when the regression was controlled for racial/ethnic and socioeconomic covariates. The percentage of Black residents in the SCA was the only covariate with a statistically significant association according to the regression coefficient (beta = 0.3892; *p*<0.0001). The results can be separately analyzed for each community detection algorithm (Supplemental Table 3), and the Stochastic Block Model uncovered SCAs with localization index anomalously lower and was not considered in the final analysis.

**Figure 1.**
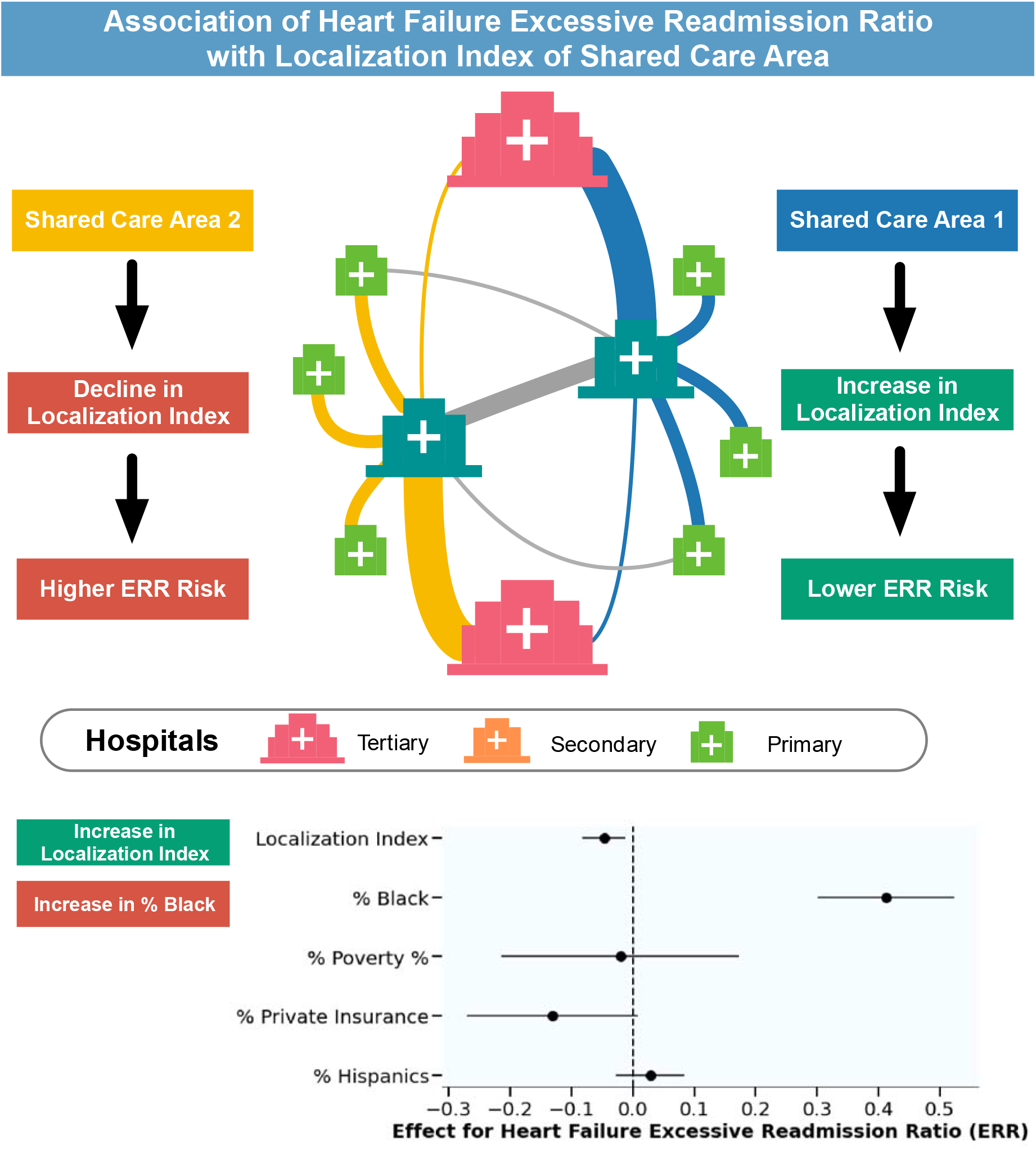
Central Illustration – Association of Heart Failure Excessive Readmission with Shared Care Networks. Hospitals are embedded in shared care areas (SCAs), which are data-driven units of care coordination emerging from the discharge networks among hospitals. The localization index (LI) is the proportion of patient discharges from hospitals within the same SCA in which these patients live. The Heart Failure Excessive Readmission Ratios (ERRs) of hospitals are associated with the SCA localization index in which they are embedded.

**Table 2.**
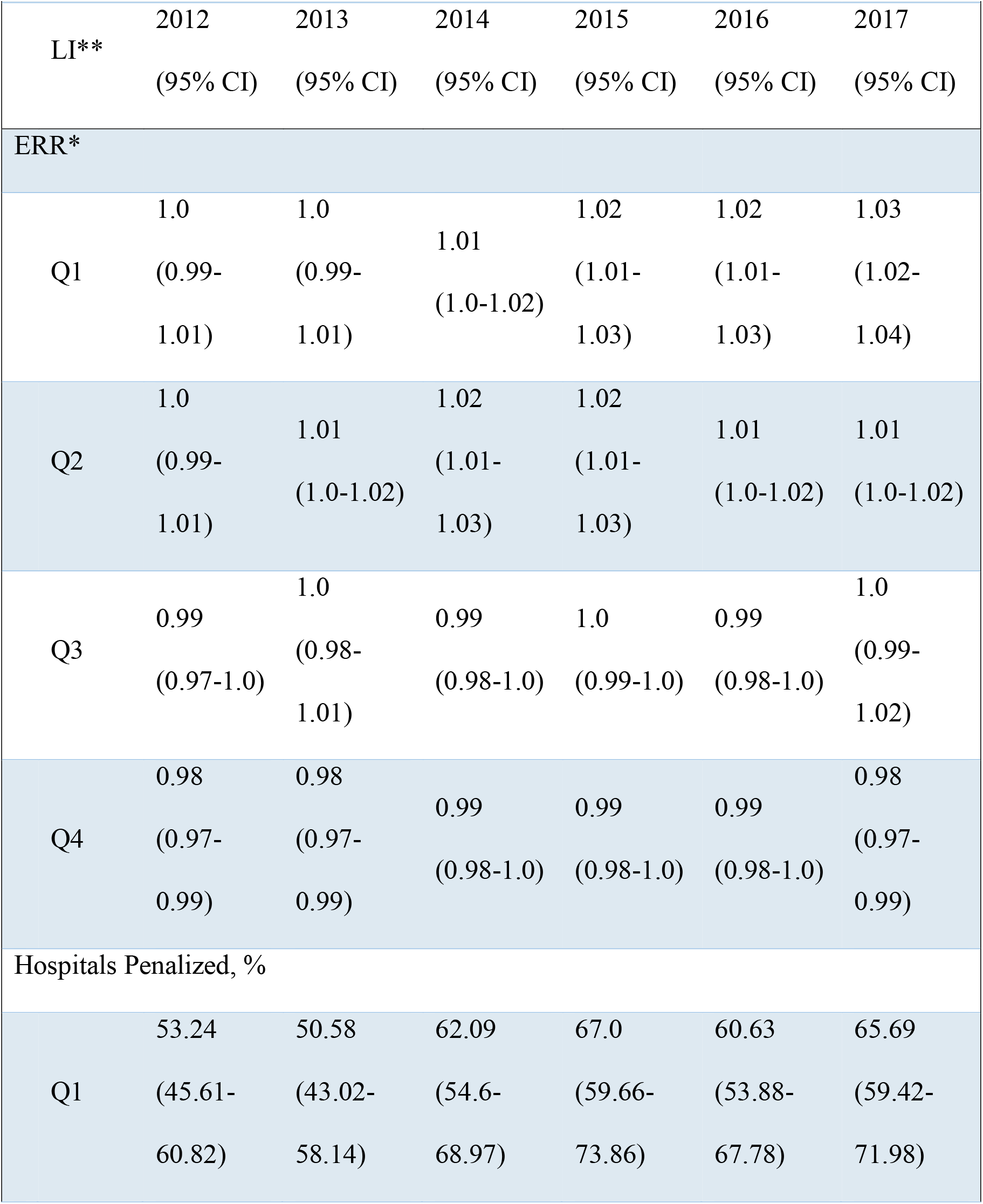
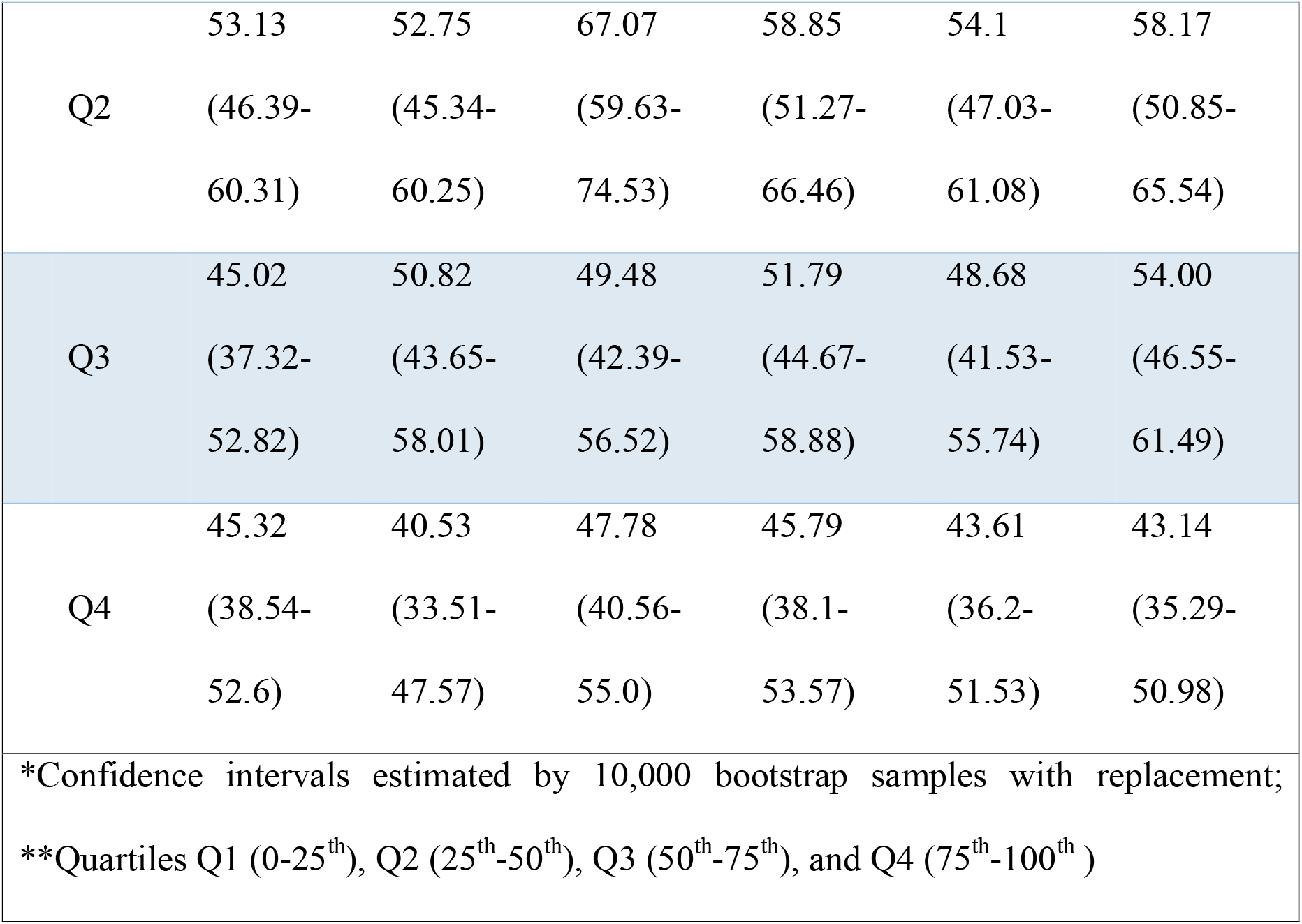
Excessive Readmission Ratios (ERR) for Hospitals in California by Localization Index (LI) Quartile.

The results of the quartile analysis indicate that the ERR of hospitals was negatively associated with the localization index (Table 3). In 2017, for instance, the ERR of hospitals in SCAs with the lowest quartile (Q1) of localization index was 1.03 (95% CI, 1.02 to 1.04) with 65.7% (95% CI, 59.4% to 72.0%) hospitals penalized. In SCAs with the highest quartile (Q4) of localization index, however, the median ERR was 0.98 (95% CI, 0.97 to 0.99) with only 43.1% (95% CI, 35.3% to 51.0%) hospitals penalized. From 2012 to 2017, the disparities between the ERR and percentage of hospitals penalized among SCAs belonging to the lowest (Q1) and highest LI (Q4) quartiles has increased mainly because of increases in the ERR and percentage of hospitals penalized hospitals within SCAs in the lowest LI quartile (Q1). In 2017, for instance, the ERR of hospitals in SCAs with the lowest quartile (Q1) of Black residents was 0.99 (95% CI, 0.98 to 1.0) with 45.2% (95% CI, 38.2% to 52.2%) hospitals penalized. In SCAs with the highest percentage of Black residents quartile (Q4), however, the median ERR was 1.03 (95% CI, 1.02 to 1.04) with 67.6% (95% CI, 60.7 to 74.6%) hospitals penalized. The percentage of Black residents is slightly higher in SCAs with lower localization (Supplemental Table 4). The results can be separately analyzed for each community detection algorithm for ERR (Supplemental Table 5), percentage of hospital penalized (Supplemental Table 6), and the percentage of Black residents (Supplemental Table 7).

**Table 3.**
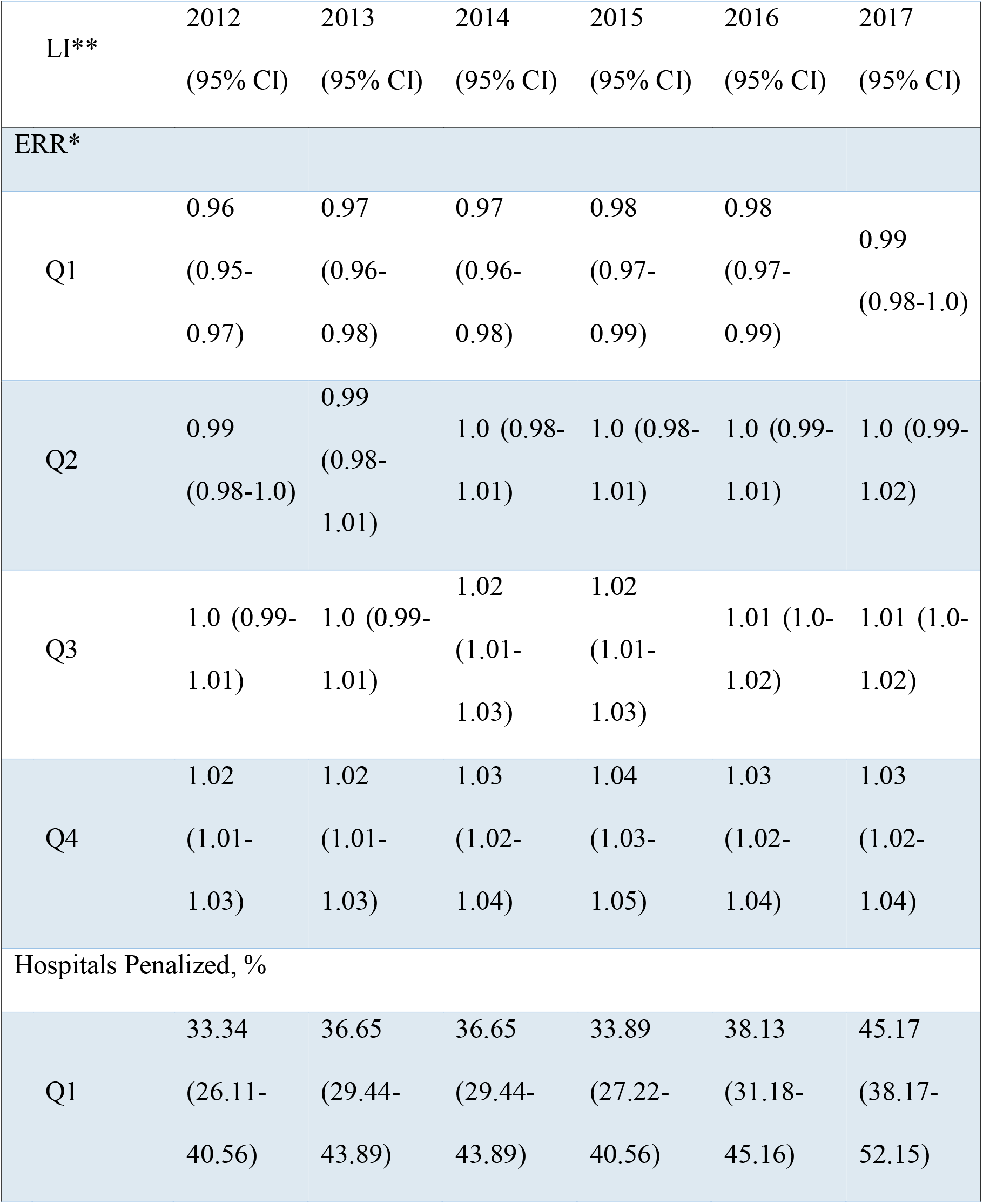
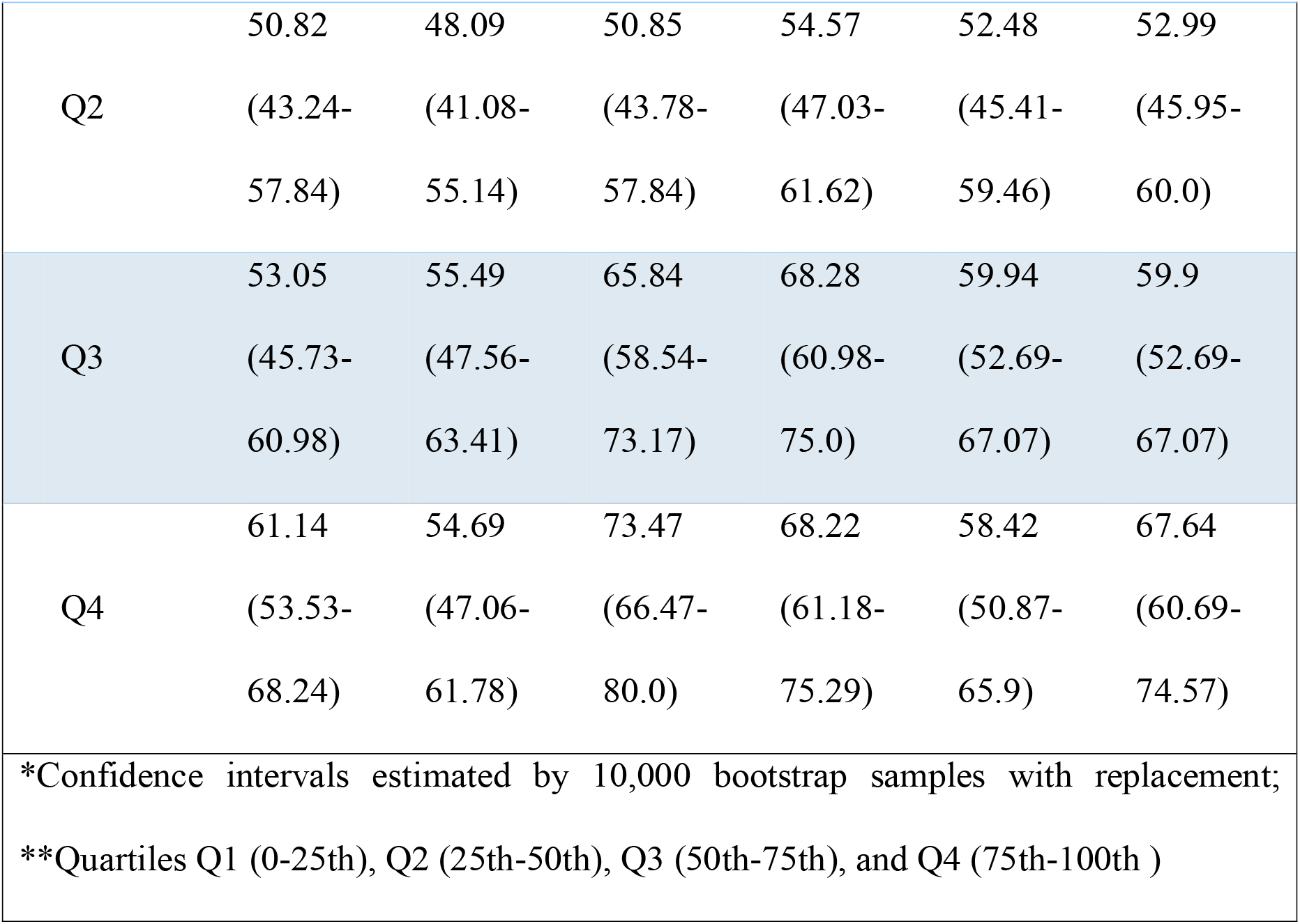
Excessive Readmission Ratios (ERR) for Hospitals in California by Percentage of Black Residents in the Shared Care Area (SCA)

**Table 4.** Results of the Generalized Estimating Equations (GEE) Regression for Excessive

| Table 4 Results of the Generalized Estimating Equations (GEE) Regression for Excessive Readmission Ratio (ERRs). |  |  |  |  |
| --- | --- | --- | --- | --- |
| Estimator | Coefficient | SE | z | P value |
| Unadjusted Model |  |  |  |  |
| Intercept | 1.0733 | 0.014 | 75.626 | <0.0001 |
| Localization Index | -0.0722 | 0.0170 | -4.2190 | <0.0001 |
| Adjusted Model |  |  |  |  |
| Intercept | 1.1054 | 0.067 | 16.558 | <0.0001 |
| Localization Index | -0.0474 | 0.0180 | -2.6670 | 0.0080 |
| % Black | 0.4128 | 0.0570 | 7.2970 | <0.0001 |
| % Poverty | -0.0208 | 0.0990 | -0.2100 | 0.8330 |
| % Private Insurance | -0.1317 | 0.0710 | -1.8500 | 0.0640 |
| % Hispanic | 0.0278 | 0.0290 | 0.9710 | 0.3320 |
| GEE using a Gaussian family and an Exchangeable working correlation structure. |  |  |  |  |

### Predictions of Excessive Readmission Ratio based on Changes in Localization Index

The predictions of ERRs and percentage of hospitals penalized based on changes in the LI (Table 5 and Figure 2) demonstrated the negative association with the LI of their SCAs as well as the positive association with the percentage of Black residents in the SCAs. The percentage range of Black residents in the stratified SCAs were 0.20% to 1.96% in Q1, 1.96% to 4.16% in Q2, 4.16% to 7.85% in Q3, and 7.85% to 17.6% in Q4. The quartiles in LI for negative differences were -0.167 (−q3), -0.058 (−q2), -0.015 (−q1); positive differences were 0.019 (+q1), 0.070 (+q2), 0.179 (+q3). In Q1 and Q4, the estimated median ERR was 0.995 (95% CI, 0.994 to 0.996) and 1.039 (95% CI, 1.038 to 1.041), respectively, with 27.5% (95% CI, 24.6% to 30.4%) and 100% (95% CI, 100% to 100%) hospitals penalized. If LI decreases by -0.167 (i.e., a *-q3* LI change), the median ERR is predicted 1.003 (9% CI, 1.002 to 1.004) and 1.047 (95% CI, 1.046 to 1.048) in Q1 and Q4, respectively, with 39.2% (95% CI, 35.8% to 42.4%) and 100% (95% CI, 100% to 100%) hospitals penalized. Conversely, if LI increases by 0.179 (i.e., a *+q4* LI change), the median ERR is predicted 0.987 (95% CI, 0.986 to 0.988) and 1.031 (95% CI, 1.030 to 1.032) in Q1 and Q4, respectively, with 18.1% (95% CI, 15.6% to 20.8%) and 91.6% (95% CI, 89.7% to 93.4%) hospitals penalized.

**Table 5.**
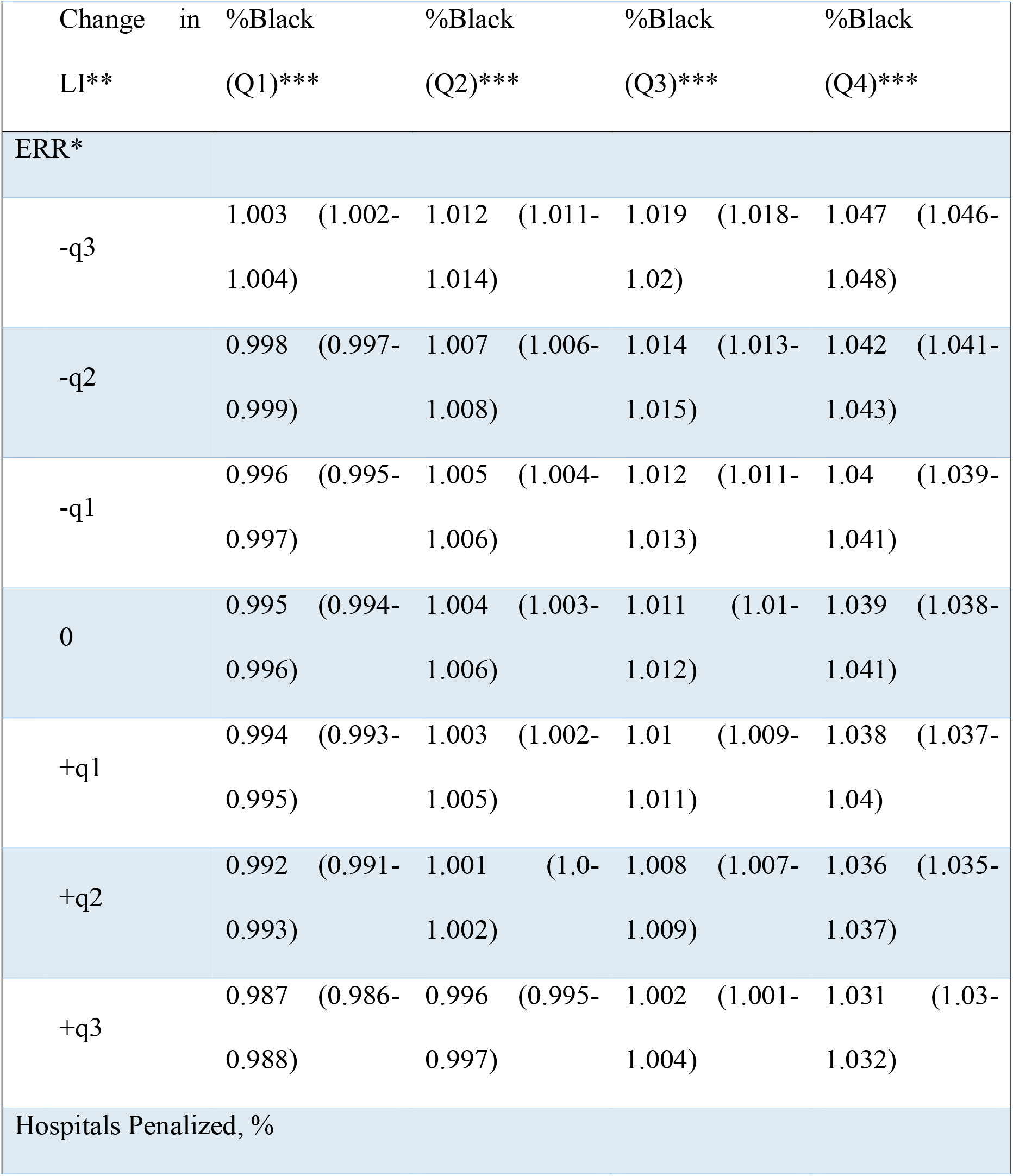
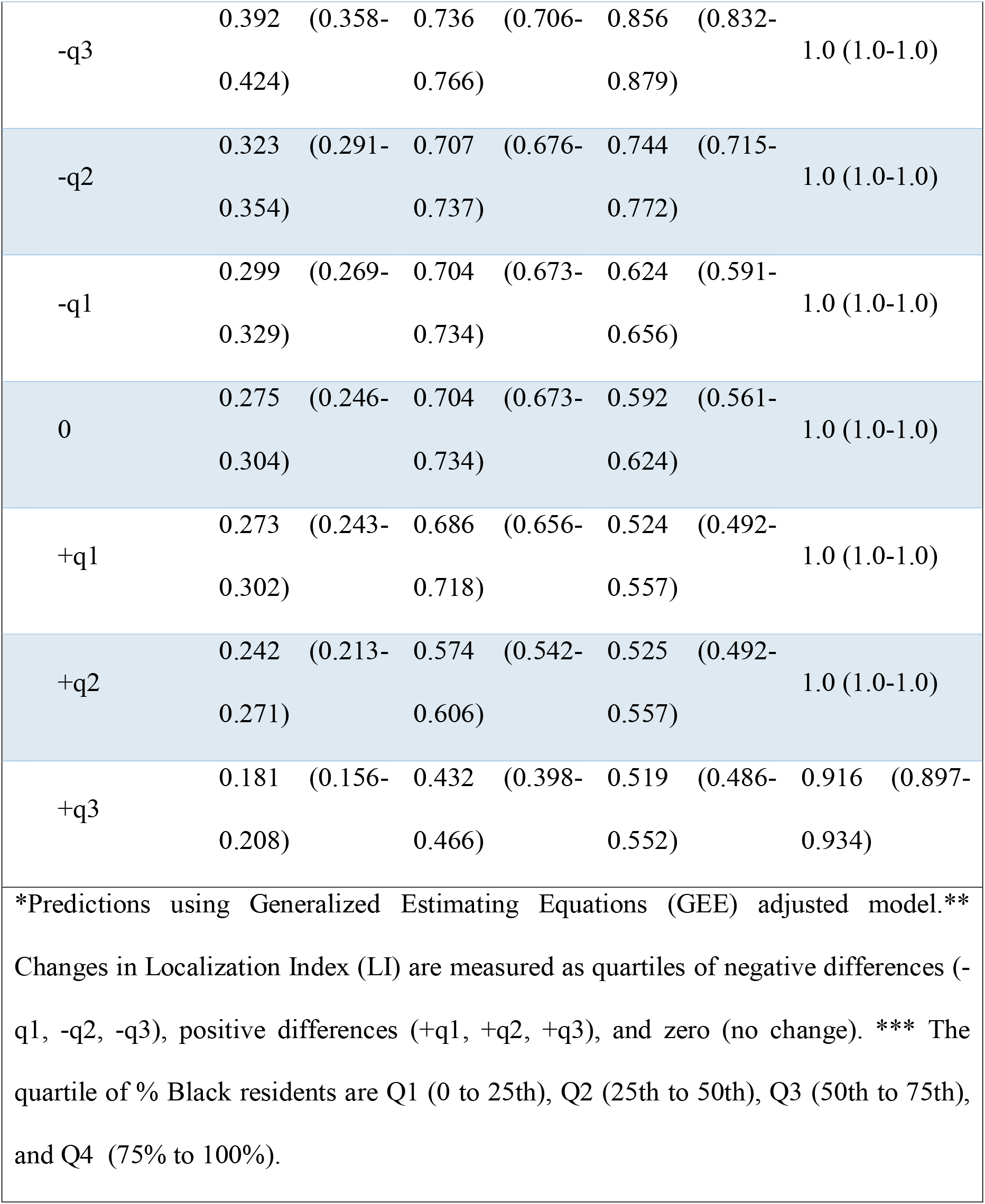
Predictions* of ERR and Percentage of Hospitals Penalized based on Changes in Localization Index (LI).

**Figure 2.**
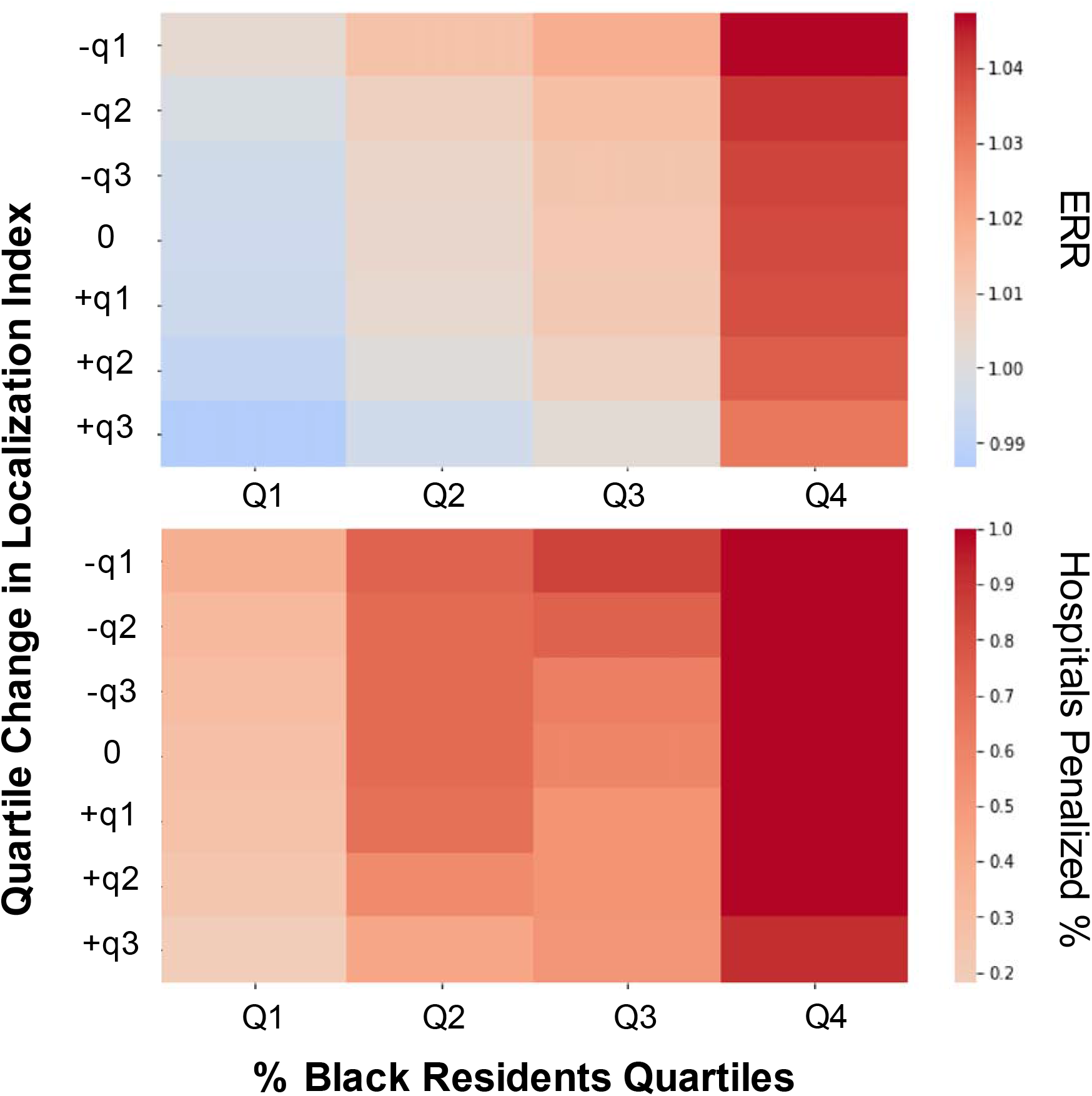
Predictions of Excessive Readmission Ratios (ERR) and Percentage of Hospitals Penalized based on Changes in Localization Index (LI) The HF excessive readmission ratios (ERRs) of hospitals are negatively associated with the localization index of the shared care areas (SCAs) in which they are embedded and positively associated with the percentage of Black residents within the SCA. The percentage of Black residents in (SCAs) were stratified into four quartiles Q1 = 0.20% to 1.96%, Q2 = 1.96% to 4.16%, Q3=4.16% to 7.85%, Q4= 7.85% to 17.6%. The quartiles in localization index differences were separately calculated for negative (−q1, -q2, -q3)=(−0.167, -0.058, and -0.015) and positive (+q1, +q2, +q3)=(0.019, 0.070, 0.179) of localization index differences.

## DISCUSSION

Regional variation in healthcare delivery is a ubiquitous phenomenon (4)(26), and the HRRP may have differently impacted almost 3 thousand US hospitals depending on their state. The main finding in the present study is that higher-than-expected HF hospital readmissions are associated with the share care networks in which hospitals are embedded. Specifically, hospitals within shared care areas (SCAs) with high localization index (LI) are associated with lower excessive readmission ratios (ERRs) than hospitals within SCAs with lower LI. The LI represents the proportion of patient discharges from hospitals within the same SCA of which these patients live. The LI is widely used as a measure of care coordination and unwarranted healthcare variation (4)(26) but to our knowledge, this is the first documentation of its association with HF higher-than-expected readmissions. In this work, the LI is ultimately derived from the shared care discharge networks. In SCAs with high LI, discharges are localized with a lower proportion of discharges of patients from other SCAs. Not only has shared care been advocated as an appropriate model to organize HF care (10)(11), but partnerships among community physicians and local hospitals have been identified as hospital strategies to reduce 30-day HF readmission (32). Characterizing shared care networks provides a roadmap for hospitals to work together, improving their shared care network as a whole instead of focusing on their hospital penalties.

Though the HRRP is a nationwide effort to reduce higher-than-expected hospital readmissions, it has also created unintended consequences in the complex system of HF care by penalizing hospitals for issues beyond their control, leaving them without specific guidance on how to improve, and focusing on punishment instead of process improvements (8). HF patients should be managed as a continuum of care within the primary, secondary and tertiary level of care, promote timely patient referrals, and care delivered within a strong working relationship (10). Integrated HF care will improve care coordination that influence patient outcomes. Features identified to result in improved shared care includes liaisons between levels of care and institutions, shared professional education, and medication optimization. Comprehensive pathways across primary, secondary, tertiary care and institutions should be developed and implemented, taking into account patients and health care providers in the design of these pathways (33).

The association of ERRs with shared care networks, however, seems to vary depending on the ethnic/racial and socioeconomic composition of SCAs. In this study, ERR is positively associated with the percentage of Blacks residents in the SCA. Ethnic/racial disparities may contribute to HF hospital readmissions (27)(29)(32)(34), and HF readmission rates are consistently higher for Black patients (34)(35)(36). In a previous case-control study (29), after matching maximum penalty hospitals as cases to their respective nearest non-penalty hospitals as controls, the authors found that maximum penalty hospitals were more likely than controls to be located in counties with low socioeconomic status.

The regional variation on the impact of the HRRP raises the following question: how much HF higher-than-expected readmissions are related to hospital-specific performance, and how much it is related to issues beyond the control of a hospital? Also, the increased association of ERR with LI in SCAs with increasingly higher percentages of Black residents raises the following question: how can improved shared care networks can reduce HF disparities among underserved and marginalized groups? Our findings will hopefully motivate cluster randomized clinical trials (37) to evaluate how improved shared care models will reduce hospital readmissions and overall costs, increase adherence to guideline-directed medical therapy, and improve clinical outcomes such as survival and development of chronic conditions.

### Study Limitations

The HRRP is a nationwide program, but our study only considered hospitals in California because large-scale hospital-specific discharge data at the ZCTA level is not publicly available to examine all US hospitals. Our finding only applies to higher-than-expected HF readmissions, and the generalization to conditions other than HF (e.g., acute myocardial infarction, pneumonia, chronic obstructive pulmonary disease) will require further investigation. The primary outcome used in our study, the ERR, is a ratio between two hospital-level regressions that can be used across heterogeneous hospitals but has little inherent variability. In its current version, our study currently neglects to model the interactions between SCAs, which deserves further investigation. While our study assumes that the ERR can be used to compare different hospitals as it accounts for a plethora of factors associated with the hospital-level HF readmissions at the individual-level, our findings should be interpreted at the hospital-level.

## Conclusions

In this study, we evaluated the association of higher-than-expected HF readmissions with shared care networks by curating publicly available large-scale hospital-level data on HF ERRs from Medicare HRRP as well as hospital-patient discharges from OSHPD. HF ERRs of hospitals were associated with the LI of the SCAs in which they were embedded, even after controlling for socioeconomic disparities. The HRRP, health systems, and hospitals should characterize and reward models of shared care practices for promoting the necessary integration capable of producing a sustainable and equitable HF care system.

## PERSPECTIVES

### Competency in Medical Knowledge

The higher-than-expected HF readmission of hospitals was associated with the shared care networks in which hospitals were embedded and the ethnic/racial composition of their shared care areas. Hospitals should collectively work to systematically improve their shared care networks for improved HF care.

### Translational Outlook

Shared care models have been advocated for HF care but have not been explicitly characterized and rewarded by nationwide control programs such as the HRRP or health systems. Improved shared care networks of HF care could mitigate higher-than-expected HF readmissions, especially among underserved and marginalized groups and translate into economic benefits. Implementation of this model will require collaboration between providers and hospital administrations. Future clinical trials will be needed to evaluate the impact of systematic implementation of improved shared care models of HF to improve higher-than-expected HF readmissions.

## Data Availability

All data used in this work is made publicly available by the Hospital Reduction Readmission Program (HRRP) and Office of Statewide Health Planning and Development (OSHPD).

https://www.cms.gov/Medicare/Medicare-Fee-for-Service-Payment/AcuteInpatientPPS/Readmissions-Reduction-Program

## Abbreviations and Acronyms

ERR: Excess Readmission Ratio
FY: Fiscal Year
GEE: Generalized Estimating Equation
HRRP: Hospital Readmission Reduction Program
LI: Localization Index
SCA: Shared Care Area
ZCTA: ZIP (Code) Tabulation Area

## Footnotes

1 https://osf.io/ckz85/

## References

(1) Papanicolas, I., Woskie, L. R., & Jha, A. K. (2018). Health Care Spending in the United States and Other High-Income Countries. Jama, 319(8), 1024–16.

(2) Benjamin, E. J., Muntner, P., Alonso, A., Bittencourt, M. S., Callaway, C. W., Carson, A. P., et al. (2019). Heart Disease and Stroke Statistics—2019 Update: A Report From the American Heart Association. Circulation, 139(10), 1.

(3) Yin, M. Y., Strege, J., Gilbert, E. M., Stehlik, J., McKellar, S. H., Elmer, A., et al. (2020). Impact of Shared Care in Remote Areas for Patients With Left Ventricular Assist Devices. J Am Coll Cardiol HF, 8(4), 302–312.

(4) Wennberg, J. E. (2020). Tracking Medicine (pp. 1–340). New York, USA: Oxford University Press.

(5) Lipsitz, L. A. (2012). Understanding Health Care as a Complex System. Jama, 308(3), 243–2.

(6) Sturmberg, J., & Lanham, given-names>H. J. (2014). Understanding health care delivery as a complex system. J Eval Clin Pract, 20(6), 1005–1009.

(7) Centers for Medicare & Medicaid Services (CMS): Hospital Readmissions Reduction Program (HRRP). Available at: https://www.cms.gov/Medicare/Medicare-Fee-for-Service-Payment/AcuteInpatientPPS/Readmissions-Reduction-Program. Accessed Nov 20, 2020

(8) Psotka, M. A., Fonarow, G. C., Allen, L. A., Maddox, K. E. J., Fiuzat, M., Heidenreich, P., et al. (2020). The Hospital Readmissions Reduction Program. J Am Coll Cardiol HF, 8(1), 1–11.

(9) Hickman, M., Drummond, N., & Grimshaw, J. (1994). A taxonomy of shared care for chronic disease. J Public Health, 16(4), 447–454.

(10) Crespo-Leiro, M. G., Metra, M., Lund, L. H., Milicic, D., Costanzo, M. R., Filippatos, G., et al. (2018). Advanced heart failure: a position statement of the Heart Failure Association of the European Society of Cardiology. Eur J Heart Fail, 20(11), 1505–1535.

(11) Huitema, A. A., Harkness, K., Heckman G. A., & McKelvie R. S. (2018). The Spoke-Hub-and-Node Model of Integrated Heart Failure Care. Can J Cardiol, 34(7), 863–870.

(12) Smith, S., Allwright, S., & ODowd, T. (2008). Does Sharing Care Across the Primary–Specialty Interface Improve Outcomes in Chronic Disease? A Systematic Review. Am J Manag Care, 213–224.

(13) Pearl, A. (2003). The effect of an integrated care approach for heart failure on general practice. Family Practice, 20(6), 642–645.

(14) Kiernan, M. S., Joseph, S. M., Katz, J. N., Kilic, A., Rich, J. D., Tallman, M. P., et al. (2015). Sharing the Care of Mechanical Circulatory Support. Circ Heart Fail, 8(3), 629–635.

(15) Hu, Y., Wang, F., & Xierali I. M. (2018). Automated Delineation of Hospital Service Areas and Hospital Referral Regions by Modularity Optimization. Health Serv Res, 53(1), 236–255.

(16) Pinheiro, D., Romero, E., & Cadeiras, M. (2019). Shared Care Areas of Heart Failure. J Card Fail, 25(8), S107–S108.

(17) Office of Statewide Health Planning and Development (OSHPD). Patient Origin/Market Share. Available at: https://data.chhs.ca.gov/dataset/patient-origin-market-share-pivot-profile-inpatient-emergency-department-and-ambulatory-surgery. Accessed August 25, 2020.

(18) Centers for Medicare & Medicaid Services (CMS). Readmission Measures Methodology, 2020 Condition-Specific Readmission Measures Updates and Specifications Report. Available at: https://qualitynet.cms.gov/inpatient/measures/readmission/methodology Accessed November 20, 2020.

(19) Uniform Data System (UDS). Zip Code to ZCTA Crosswalk. Available at: https://udsmapper.org/zip-code-to-zcta-crosswalk Accessed November 20, 2020.

(20) Pinheiro, D., Hartman, R., Romero, E., Menezes, R., & Cadeiras, M. (2020). Network-Based Delineation of Health Service Areas: A Comparative Analysis of Community Detection Algorithms. In Complex Netw XI (Vol. 2008, pp. 359–370). Cham: Springer International Publishing.

(21) Blondel V. D., Guillaume, J.-L., Lambiotte, R., & Lefebvre, E. (2008). Fast unfolding of communities in large networks. J Stat Mech, 2008(10).

(22) Peixoto, T. P. (2014). Hierarchical Block Structures and High-Resolution Model Selection in Large Networks. Phys Rev X, 4(1), 9851042–18.

(23) Peixoto, T. P. (2015). Model Selection and Hypothesis Testing for Large-Scale Network Models with Overlapping Groups. Phys Rev X, 5(1), 1981–20.

(24) Rosvall, M., & Bergstrom C. T. (2008). Maps of random walks on complex networks reveal community structure. Proc Natl Acad Sci U S A, 105(4), 1118–1123.

(25) Xie, J., Szymanski, B. K., & Liu, X. (2011). SLPA: Uncovering Overlapping Communities in Social Networks via a Speaker-Listener Interaction Dynamic Process. 2011 IEEE 11th IEEE Int Conf Data Min Workshops, 344–349.

(26) Wennberg, J. (1996). The Dartmouth Atlas of Health Care. (M. M. Cooper, Ed.). Chicago, IL: American Hospital Association.

(27) Wallace, J., Mohan, D., Angus, D. C., Driessen, J. R., Seymour, C. M., Yealy, D. M., et al. (2018). Referral Regions for Time-Sensitive Acute Care Conditions in the United States. Ann Emerg Med, 72(2), 1–9.

(28) Liang K.-Y., & Zeger S. L. (1986). Longitudinal data analysis using generalized linear models. Biometrika, 73(1), 13–22.

(29) Caracciolo, C., Parker, D., Marshall, E., & Brown, J. (2017). Excess Readmission vs Excess Penalties: Maximum Readmission Penalties as a Function of Socioeconomic and Geography. J Hosp Med, 12(8), 610–617.

(30) Python Package Index. Statsmodels. Available at: https://pypi.org/project/statsmodels/. Accessed August 25, 2020.

(31) Python Package Index. Bootstrapped. Available at: https://pypi.org/project/bootstrapped/. Accessed August 25, 2020.

(32) Bradley, E. H., Curry, L., Horwitz, L. I., Sipsma, H., Wang, Y., Walsh, M. N., et al. (2013). Hospital Strategies Associated With 30-Day Readmission Rates for Patients With Heart Failure. Circ Cardiovasc Qual Outcomes, 6(4), 444–450.

(33) MacInnes, J., & Williams, L. (2018). A review of integrated heart failure care. Primary Health Care Research & Development, 20, 1–8.

(34) Chaiyachati, K. H., Qi, M., & Werner R. M. (2018). Changes to Racial Disparities in Readmission Rates After Medicare’s Hospital Readmissions Reduction Program Within Safety-Net and Non–Safety-Net Hospitals. JAMA Netw Open, 1(7), e184154–12.

(35) Durstenfeld, M. S., Ogedegbe, O., Katz, S. D., Park, H., & Blecker, S. (2016). Racial and Ethnic Differences in Heart Failure Readmissions and Mortality in a Large Municipal Healthcare System. J Am Coll Cardiol HF, 4(11), 885–893.

(36) Churchwell, K., Elkind M. S. V., Benjamin R. M., Carson A. P., Chang E. K., Lawrence, W., et al. (2020). Call to Action: Structural Racism as a Fundamental Driver of Health Disparities: A Presidential Advisory From the American Heart Association. Circulation,

(37) Mercer, T., Njuguna, B., Bloomfield, G. S., Dick, J., Finkelstein, E., Kamano, J., et al. (2019). Strengthening Referral Networks for Management of Hypertension Across the Health System (STRENGTHS) in western Kenya: a study protocol of a cluster randomized trial. Trials, 20(1), 1–13.

