## Supplementary Material for "Association of Shared Care Networks with Heart Failure Excessive Hospital Readmissions"

^c^Independent Researcher

^d^Polytechnic School of Pernambuco, University of Pernambuco, Recife, Brazil

^e^Division of Cardiovascular Surgery, University of Pernambuco, Recife, Brazil

^f^Family Caregiving Institute at the Betty Irene Moore School of Nursing; University of California, Davis

^g^Department of Public Health Sciences, Division of Biostatistics, University of California, Davis, Davis, USA

### 1. Supplemental Methods

#### 1.1 Measuring Higher-Than-Expected Heart Failure Readmissions

Higher-than-expected HF readmission were measured as the Excessive Readmission Ratio (ERR) was used to measure. The ERR methodology is accessible in the Condition-Specific Readmission Measures Updates and Specifications Report from the Centers for Medicare & Medicaid Services (CMS) (1). This ERR data is publicly available from the Hospital Reduction Readmission Program (HRRP) (2). It is important to point out that the ERR data of each year in the period from 2012 to 2017 (i.e., Fiscal Year [FY] 2014 and 2019) was separately downloaded and compiled into a single file available at the repository of this project (Supplemental Table S1).

#### 1.2 Uncovering Shared Care Areas from Hospital-Patient Discharge Data

Shared Care Areas (SCAs) were delineated using algorithms of community detection. A minimally sufficient set of four diverse community detection algorithms were considered to decrease both variability and bias (3) (Table S2 in Supplement Material). The Stochastic Block Model algorithm was removed from the analysis because it delineates SCAs with localization index as lower as 0.3 (95% CI 0.23 to 0.37).

### 2. Supplemental Tables

| Supplemental Table 1 Excessive Readmission Ratios data files obtained from Centers for Medicare & Medicaid Services (CMS) - Hospital Readmissions Reduction Program (HRRP). | | | |
| --- | --- | --- | --- |
| Year | Filename | Sheet Name | Header |
| 2014 | FY 2014 Final Rule Readmissions Supplemental Data PUF-CN_Sept 2013.xlsx | Final FY 2014-CN2-Sept 2013 | 2 |
| 2015 | FY 2015 IPPS Final Rule Readmissions PUF-Oct 2014 CN.xls | Final FY15-CN Oct 2014 | 1 |
| 2016 | FY 2016 IPPS Final Rule Readmissions PUF revised 08-04-15.xlsx | Final Rule FY 2016 | 1 |
| 2017 | FY 2017 IPPS Final Rule Readmissions Supplemental Data File.xlsx | Final Rule FY 2017 | 1 |
| 2018 | FY 2018 IPPS Final Rule Readmissions Supplemental Data File.xlsx | FR FY 2018 | 1 |
| 2019 | fy_2019_ipps_final_rule_hrrp_supplemental_file.xlsx | FR FY 2019 | 1 |

| Supplemental Table 2 Algorithms of community detection used in the delineation of Shared Care Areas (SCAs). | |  |  |
| --- | --- | --- | --- |
| Algorithm | Description | Parameters | Reference |
| Louvain | Maximizes the network modularity, which is the extent to which the density of links within the found communities excessively surpasses what would be expected if links were placed at randomly. | resolution=1.0 | (4) |
| Stochastic Block Model | Infers the block structure of the network using a maximum likelihood estimator. | Degree-corrected = True. | (5-6) |
| Infomap | Maximizes the map equation, which is the length of the coding scheme necessary to communicate the sequence of movements of random walkers within the network. | Algorithm = two-level Trials=30 | (7) |
| Speaker-Listener Label Propagation Algorithm (SLPA). | Propagates the most common labels among neighbors until the formation of consensus groups. | Post processing threshold (r) = 0.5 | [8] |

| Supplemental Table 3 Localization Index (LI) of Shared Care Areas (SCA) found by Community Detection Algorithms. | | | | | | | |
| --- | --- | --- | --- | --- | --- | --- | --- |
|  | LI** | 2012 (95% CI) | 2013 (95% CI) | 2014 (95% CI) | 2015 (95% CI) | 2016 (95% CI) | 2017 (95% CI) |
| Blockmodel* | |  |  |  |  |  |  |
|  | 0-25 | 0.3  (0.23-0.37) | 0.3  (0.22-0.37) | 0.32  (0.25-0.39) | 0.4  (0.34-0.45) | 0.41  (0.36-0.46) | 0.41 (0.35-0.46) |
|  | 25-50 | 0.68  (0.67-0.68) | 0.67  (0.66-0.69) | 0.68  (0.67-0.69) | 0.7  (0.69-0.71) | 0.64  (0.64-0.64) | 0.67  (0.66-0.69) |
|  | 50-75 | 0.87  (0.86-0.89) | 0.87  (0.86-0.88) | 0.83  (0.83-0.84) | 0.86  (0.85-0.88) | 0.83  (0.81-0.84) | 0.84  (0.82-0.85) |
|  | 75-100 | 0.95  (0.94-0.95) | 0.95  (0.94-0.95) | 0.94  (0.93-0.94) | 0.94  (0.94-0.95) | 0.95  (0.94-0.95) | 0.95  (0.94-0.95) |
| Infomap* | |  |  |  |  |  |  |
|  | 0-25 | 0.71  (0.69-0.73) | 0.71  (0.69-0.73) | 0.71  (0.69-0.73) | 0.71  (0.69-0.73) | 0.71  (0.69-0.73) | 0.71  (0.7-0.73) |
|  | 25-50 | 0.87  (0.86-0.87) | 0.86  (0.85-0.86) | 0.86  (0.85-0.86) | 0.85  (0.84-0.86) | 0.85  (0.84-0.86) | 0.86  (0.85-0.87) |
|  | 50-75 | 0.92  (0.91-0.92) | 0.91  (0.91-0.92) | 0.91  (0.91-0.92) | 0.91  (0.91-0.92) | 0.91  (0.91-0.92) | 0.92  (0.91-0.92) |
|  | 75-100 | 0.95  (0.95-0.95) | 0.95  (0.95-0.95) | 0.95  (0.95-0.95) | 0.95  (0.95-0.95) | 0.95  (0.95-0.95) | 0.95  (0.95-0.95) |
| Louvain* | |  |  |  |  |  |  |
|  | 0-25 | 0.85  (0.83-0.86) | 0.85  (0.83-0.86) | 0.85  (0.83-0.87) | 0.85  (0.83-0.86) | 0.84  (0.83-0.86) | 0.84  (0.83-0.86) |
|  | 25-50 | 0.91  (0.91-0.91) | 0.91  (0.91-0.91) | 0.91  (0.91-0.91) | 0.91  (0.91-0.91) | 0.91  (0.91-0.91) | 0.91  (0.91-0.91) |
|  | 50-75 | 0.92  (0.92-0.92) | 0.93  (0.92-0.93) | 0.93  (0.93-0.93) | 0.93  (0.92-0.93) | 0.93  (0.93-0.93) | 0.93  (0.92-0.93) |
|  | 75-100 | 0.96  (0.95-0.96) | 0.96  (0.96-0.96) | 0.96  (0.96-0.96) | 0.96  (0.95-0.96) | 0.95  (0.95-0.96) | 0.95  (0.95-0.96) |
| SLPA* | |  |  |  |  |  |  |
|  | 0-25 | 0.54  (0.51-0.57) | 0.55  (0.53-0.57) | 0.56  (0.53-0.58) | 0.55  (0.53-0.57) | 0.55  (0.53-0.57) | 0.58  (0.56-0.59) |
|  | 25-50 | 0.69  (0.68-0.7) | 0.7  (0.69-0.7) | 0.7  (0.69-0.71) | 0.7 (0.69-0.71) | 0.69  (0.68-0.7) | 0.71  (0.7-0.72) |
|  | 50-75 | 0.82  (0.81-0.82) | 0.82  (0.81-0.83) | 0.83  (0.82-0.83) | 0.83  (0.82-0.84) | 0.83  (0.82-0.83) | 0.83  (0.82-0.84) |
|  | 75-100 | 0.91  (0.9-0.92) | 0.91  (0.9-0.92) | 0.91  (0.91-0.92) | 0.91  (0.9-0.92) | 0.91  (0.9-0.92) | 0.91  (0.91-0.92) |
| *Confidence intervals estimated by bootstrapping with replacement; **Percentile ranges | | | | | | | |

| Supplemental Table 4 Percentage of Black Residents within SCA by Quartiles | | | | | |  |  |
| --- | --- | --- | --- | --- | --- | --- | --- |
|  | %Black** | 2012 (95% CI) | 2013 (95% CI) | 2014 (95% CI) | 2015 (95% CI) | 2016 (95% CI) | 2017 (95% CI) |
| %Black* | |  |  |  |  |  |  |
|  | Q1 | 1.53 (1.47-1.58) | 1.53 (1.47-1.59) | 1.53 (1.47-1.59) | 1.53 (1.47-1.59) | 1.53 (1.48-1.59) | 1.53 (1.48-1.59) |
|  | Q2 | 3.11 (2.98-3.24) | 3.11 (2.99-3.24) | 3.11 (2.98-3.24) | 3.11 (2.98-3.24) | 3.11 (2.98-3.24) | 3.11 (2.98-3.24) |
|  | Q3 | 5.78 (5.59-5.96) | 5.78 (5.59-5.96) | 5.78 (5.59-5.96) | 5.78 (5.59-5.96) | 5.79 (5.61-5.97) | 5.79 (5.61-5.98) |
|  | Q4 | 11.0 (10.73-11.28) | 11.0 (10.73-11.28) | 11.0 (10.74-11.28) | 11.0 (10.73-11.29) | 11.04 (10.77-11.33) | 11.04 (10.76-11.33) |
| *Confidence intervals estimated by 10,000 bootstrap samples with replacement; **Quartiles Q1 (0-25th), Q2 (25th-50th), Q3 (50th-75th), and Q4 (75th-100th ) | | | | | | | |

| Supplemental Table 5 Excessive Readmission Ratio (ERR) for Heart Failure by Localization Index (LI) of Shared Care Areas (SCA) found by Community Detection Algorithms. | | | | | | | |
| --- | --- | --- | --- | --- | --- | --- | --- |
|  | LI** | 2012 (95% CI) | 2013 (95% CI) | 2014 (95% CI) | 2015 (95% CI) | 2016 (95% CI) | 2017 (95% CI) |
| Blockmodel* | |  |  |  |  |  |  |
|  | Q1 | 1.01 (0.99-1.03) | 1.01 (0.99-1.03) | 1.02 (1.0-1.03) | 1.02 (1.01-1.04) | 1.02 (1.01-1.04) | 1.03 (1.01-1.04) |
|  | Q2 | 0.99 (0.98-1.01) | 1.01 (0.99-1.02) | 1.01 (1.0-1.03) | 1.01 (0.99-1.02) | 1.02 (0.99-1.04) | 1.02 (1.0-1.04) |
|  | Q3 | 0.99 (0.96-1.01) | 0.99 (0.96-1.01) | 1.0 (0.97-1.03) | 0.99 (0.97-1.01) | 0.99 (0.98-1.01) | 1.0 (0.98-1.02) |
|  | Q4 | 0.98 (0.95-1.0) | 0.98 (0.96-1.0) | 0.99 (0.97-1.0) | 0.99 (0.98-1.01) | 0.99 (0.97-1.01) | 0.99 (0.97-1.01) |
| Infomap* | |  |  |  |  |  |  |
|  | Q1 | 1.01 (0.99-1.02) | 1.02 (1.0-1.03) | 1.03 (1.01-1.05) | 1.03 (1.01-1.05) | 1.03 (1.01-1.05) | 1.03 (1.01-1.05) |
|  | Q2 | 1.0 (0.98-1.02) | 1.0 (0.98-1.02) | 1.01 (0.99-1.03) | 1.02 (1.0-1.03) | 1.0 (0.98-1.02) | 1.01 (0.99-1.02) |
|  | Q3 | 0.98 (0.95-1.0) | 0.99 (0.97-1.01) | 0.99 (0.97-1.01) | 0.99 (0.98-1.01) | 1.0 (0.98-1.01) | 1.01 (0.99-1.03) |
|  | Q4 | 0.98 (0.96-1.0) | 0.98 (0.96-1.0) | 0.99 (0.97-1.01) | 0.99 (0.96-1.01) | 0.99 (0.97-1.01) | 0.99 (0.97-1.01) |
| Louvain* | |  |  |  |  |  |  |
|  | Q1 | 0.99 (0.97-1.0) | 0.99 (0.97-1.01) | 1.01 (0.99-1.03) | 1.01 (1.0-1.03) | 1.01 (1.0-1.03) | 1.02 (1.0-1.04) |
|  | Q2 | 0.99 (0.98-1.01) | 1.0 (0.98-1.02) | 1.02 (1.0-1.04) | 1.02 (1.0-1.05) | 1.02 (1.01-1.04) | 1.03 (1.01-1.04) |
|  | Q3 | 1.0 (0.98-1.02) | 1.0 (0.98-1.02) | 1.0 (0.99-1.02) | 1.0 (0.98-1.02) | 0.99 (0.97-1.01) | 1.0 (0.98-1.02) |
|  | Q4 | 0.99 (0.97-1.01) | 0.99 (0.97-1.01) | 0.99 (0.97-1.01) | 0.99 (0.97-1.01) | 0.99 (0.97-1.01) | 0.99 (0.97-1.01) |
| SLPA* | |  |  |  |  |  |  |
|  | Q1 | 0.99 (0.98-1.01) | 1.0 (0.98-1.02) | 1.01 (0.99-1.02) | 1.02 (1.01-1.04) | 1.02 (1.01-1.04) | 1.03 (1.02-1.05) |
|  | Q2 | 1.01 (1.0-1.03) | 1.01 (1.0-1.03) | 1.03 (1.02-1.05) | 1.02 (1.0-1.04) | 1.01 (0.99-1.03) | 0.99 (0.97-1.02) |
|  | Q3 | 0.98 (0.96-1.0) | 0.99 (0.97-1.02) | 0.98 (0.97-1.0) | 0.99 (0.97-1.01) | 0.99 (0.97-1.01) | 1.01 (0.99-1.03) |
|  | Q4 | 0.98 (0.96-1.0) | 0.98 (0.96-1.0) | 0.99 (0.97-1.01) | 0.99 (0.97-1.01) | 0.99 (0.97-1.02) | 0.98 (0.96-1.0) |
| *Confidence intervals estimated by 10,000 bootstrap samples with replacement; **Quartiles Q1 (0-25th), Q2 (25th-50th), Q3 (50th-75th), and Q4 (75th-100th ) | | | | | | | |

| Supplemental Table 6 Percentage of Hospital Penalized for Heart Failure Excessive Readmission Ratios by Localization Index (LI) of Shared Care Areas (SCA) and Community Detection Algorithms. | | | | | | | |
| --- | --- | --- | --- | --- | --- | --- | --- |
|  | LI** | 2012 (95% CI) | 2013 (95% CI) | 2014 (95% CI) | 2015 (95% CI) | 2016 (95% CI) | 2017 (95% CI) |
| Blockmodel* | |  |  |  |  |  |  |
|  | 0-25 | 58.28 (43.75-72.92) | 52.11 (37.5-66.67) | 65.44 (51.92-78.85) | 65.76 (54.29-77.14) | 64.35 (53.95-75.0) | 69.78 (59.21-80.26) |
|  | 25-50 | 50.81 (39.24-62.03) | 51.93 (40.51-63.29) | 54.73 (44.0-65.33) | 52.7 (40.35-64.91) | 62.57 (41.67-83.33) | 65.86 (51.43-80.0) |
|  | 50-75 | 50.96 (36.73-65.31) | 49.02 (34.69-63.27) | 61.46 (42.31-80.77) | 55.16 (40.82-69.39) | 42.58 (32.18-52.87) | 46.09 (35.53-56.58) |
|  | 75-100 | 38.61 (26.32-50.88) | 40.52 (28.07-52.63) | 49.97 (38.75-61.25) | 47.41 (35.09-59.65) | 44.02 (30.0-58.0) | 44.0 (30.0-58.0) |
| Infomap* | |  |  |  |  |  |  |
|  | 0-25 | 60.68 (49.18-72.13) | 51.61 (38.33-63.37) | 64.87 (52.63-77.19) | 64.86 (52.63-77.19) | 64.38 (52.54-76.27) | 70.94 (59.68-82.26) |
|  | 25-50 | 48.43 (35.94-60.94) | 48.97 (35.29-62.75) | 64.35 (51.79-76.79) | 65.31 (51.92-78.85) | 49.1 (36.84-61.4) | 49.26 (38.03-60.56) |
|  | 50-75 | 42.19 (28.89-57.78) | 50.83 (37.29-64.41) | 49.17 (36.36-61.82) | 49.24 (37.31-61.19) | 48.57 (37.14-60.0) | 61.87 (49.09-74.55) |
|  | 75-100 | 44.53 (33.33-57.14) | 42.95 (30.16-55.56) | 47.64 (35.38-60.0) | 45.59 (33.33-57.89) | 45.1 (31.37-58.82) | 40.9 (26.53-55.1) |
| Louvain* | |  |  |  |  |  |  |
|  | 0-25 | 46.56 (33.93-58.93) | 51.83 (39.29-64.29) | 63.38 (51.67-75.0) | 66.62 (55.0-78.33) | 56.61 (43.33-68.33) | 53.44 (40.0-66.67) |
|  | 25-50 | 49.99 (39.19-60.81) | 48.18 (34.62-61.54) | 64.49 (50.0-77.08) | 58.27 (43.75-72.92) | 59.13 (46.97-71.21) | 71.17 (60.61-81.82) |
|  | 50-75 | 56.4 (41.03-71.79) | 52.91 (41.18-64.71) | 52.96 (41.18-64.71) | 55.79 (44.12-67.65) | 49.98 (37.04-62.96) | 48.08 (35.19-61.11) |
|  | 75-100 | 47.0 (34.38-59.38) | 40.14 (28.07-52.63) | 45.59 (33.33-57.89) | 42.07 (29.82-54.39) | 40.37 (28.07-52.63) | 49.14 (36.84-63.16) |
| SLPA* |  |  |  |  |  |  |  |
|  | 0-25 | 51.82 (38.89-64.81) | 48.28 (35.71-60.71) | 57.9 (45.61-70.18) | 69.47 (57.63-81.36) | 60.71 (47.54-73.77) | 70.46 (61.18-80.0) |
|  | 25-50 | 62.48 (50.0-75.0) | 60.37 (48.28-72.41) | 71.93 (59.65-82.46) | 53.37 (41.38-65.52) | 53.25 (40.32-66.13) | 52.49 (37.5-67.5) |
|  | 50-75 | 39.64 (27.59-51.72) | 48.09 (35.19-61.11) | 45.88 (34.43-59.02) | 49.91 (37.1-62.9) | 47.45 (35.59-61.02) | 52.2 (40.0-64.62) |
|  | 75-100 | 44.72 (32.31-56.92) | 38.36 (27.69-50.77) | 49.95 (36.21-62.07) | 50.12 (37.04-62.96) | 45.42 (32.73-58.18) | 38.27 (25.53-53.19) |
| *Confidence intervals estimated by bootstrapping with replacement; **Percentile ranges | | | | | | | |

| Supplemental Table 7 Percentage of Black population by Localization Index (LI) of Shared Care Areas (SCA). | | | | | | | |
| --- | --- | --- | --- | --- | --- | --- | --- |
|  | LI** | 2012 (95% CI) | 2013 (95% CI) | 2014 (95% CI) | 2015 (95% CI) | 2016 (95% CI) | 2017 (95% CI) |
| Infomap* | |  |  |  |  |  |  |
|  | 0-25 | 6.61 (5.47-7.77) | 6.64 (5.47-7.87) | 6.73 (5.46-7.98) | 6.74 (5.48-8.01) | 6.79 (5.55-8.04) | 6.57 (5.37-7.76) |
|  | 25-50 | 6.46 (5.43-7.52) | 7.25 (6.07-8.45) | 6.94 (5.81-8.09) | 7.32 (6.2-8.5) | 6.79 (5.67-7.95) | 6.92 (6.03-7.83) |
|  | 50-75 | 3.58 (2.91-4.3) | 3.55 (3.03-4.1) | 3.54 (2.96-4.14) | 3.35 (2.88-3.86) | 3.43 (2.96-3.9) | 2.37 (2.12-2.62) |
|  | 75-100 | 3.62 (3.11-4.16) | 3.62 (3.11-4.15) | 3.68 (3.17-4.21) | 3.81 (3.25-4.38) | 3.96 (3.35-4.58) | 4.06 (3.44-4.68) |
| Louvain* | |  |  |  |  |  |  |
|  | 0-25 | 3.8 (3.15-4.59) | 3.8 (3.15-4.57) | 4.29 (3.52-5.15) | 4.29 (3.53-5.16) | 4.3 (3.53-5.15) | 4.29 (3.51-5.15) |
|  | 25-50 | 6.65 (5.73-7.56) | 8.57 (7.58-9.44) | 8.34 (7.28-9.23) | 8.35 (7.28-9.23) | 8.12 (7.33-8.82) | 8.12 (7.36-8.82) |
|  | 50-75 | 7.04 (6.13-7.93) | 5.14 (4.4-5.88) | 5.15 (4.41-5.91) | 5.15 (4.42-5.91) | 4.36 (3.54-5.25) | 4.35 (3.54-5.22) |
|  | 75-100 | 4.4 (4.0-4.8) | 4.43 (3.99-4.87) | 4.44 (3.99-4.87) | 4.43 (3.99-4.87) | 4.44 (3.98-4.88) | 4.43 (4.0-4.87) |
| SLPA* | |  |  |  |  |  |  |
|  | 0-25 | 4.57 (3.6-5.63) | 4.8 (3.86-5.85) | 4.78 (3.85-5.79) | 5.06 (4.09-6.11) | 5.27 (4.26-6.4) | 6.69 (5.75-7.63) |
|  | 25-50 | 7.13 (6.06-8.19) | 6.61 (5.5-7.7) | 6.62 (5.47-7.72) | 6.42 (5.31-7.54) | 6.27 (5.22-7.37) | 3.76 (2.91-4.77) |
|  | 50-75 | 4.47 (3.35-5.72) | 4.73 (3.55-6.06) | 4.83 (3.73-6.0) | 4.97 (3.9-6.14) | 4.94 (3.81-6.17) | 4.67 (3.63-5.8) |
|  | 75-100 | 4.56 (3.86-5.31) | 4.51 (3.81-5.26) | 4.43 (3.7-5.22) | 4.09 (3.43-4.83) | 4.11 (3.45-4.84) | 4.39 (3.64-5.22) |
| *Confidence intervals estimated by bootstrapping with replacement; **Percentile ranges | | | | | | | |

### 3. Supplemental Figures

#### Supplemental Figure S1

The Excess Readmission Ratio (ERR) of hospitals and the localization index of their Shared Care Areas (SCAs) were integrated each year by linking the ZCTA of hospitals to the set of ZCTAs of SCAs.

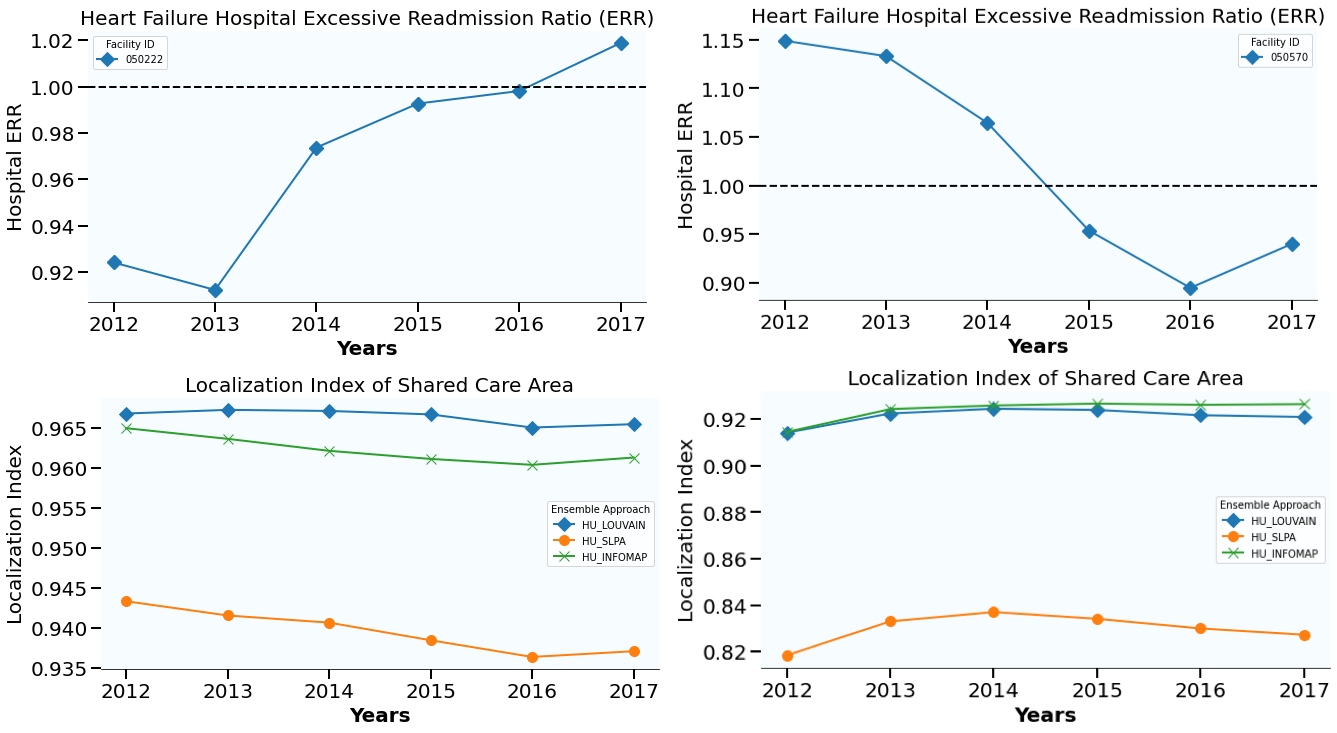

### References

(1) Centers for Medicare & Medicaid Services (CMS). Readmission Measures Methodology, 2020 Condition-Specific Readmission Measures Updates and Specifications Report. Accessed November 20, 2020. Available from: <https://qualitynet.cms.gov/inpatient/measures/readmission/methodology>

(2) Centers for Medicare & Medicaid Services (CMS): Hospital Readmissions Reduction Program (HRRP). Accessed November 20, 2020. Available from: <https://www.cms.gov/Medicare/Medicare-Fee-for-Service-Payment/AcuteInpatientPPS/Readmissions-Reduction-Program>

(3) Pinheiro, D., Hartman, R., Romero, E., Menezes, R., & Cadeiras, M. (2020). Network-Based Delineation of Health Service Areas: A Comparative Analysis of Community Detection Algorithms. In *Complex Networks XI* (Vol. 2008, pp. 359–370). Cham: Springer International Publishing. <http://doi.org/10.1007/978-3-030-40943-2_30>

(4) Blondel, V. D., Guillaume, J.-L., Lambiotte, R., & Lefebvre, E. (2008). Fast unfolding of communities in large networks. *Journal of Statistical Mechanics-Theory and Experiment*, *2008*(10). <http://doi.org/10.1088/1742-5468/2008/10/P10008>

[5] Peixoto, T. P. (2014). Hierarchical Block Structures and High-Resolution Model Selection in Large Networks. *Physical Review X*, *4*(1), 9851042–18. <http://doi.org/10.1103/PhysRevX.4.011047>

[6] Peixoto, T. P. (2015). Model Selection and Hypothesis Testing for Large-Scale Network Models with Overlapping Groups. *Physical Review X*, *5*(1), 1981–20. <http://doi.org/10.1103/PhysRevX.5.011033>

[7] Rosvall, M., & Bergstrom, C. T. (2008). Maps of random walks on complex networks reveal community structure. *Proceedings of the National Academy of Sciences of the United States of America*, *105*(4), 1118–1123. <http://doi.org/10.1073/pnas.0706851105>

(8) Xie, J., Szymanski, B. K., & Liu, X. (2011). SLPA: Uncovering Overlapping Communities in Social Networks via a Speaker-Listener Interaction Dynamic Process. *2011 IEEE 11th International Conference on Data Mining Workshops*, 344–349. <http://doi.org/10.1109/ICDMW.2011.154>
